# Real-World Effectiveness and Safety of Tocilizumab in Refractory Rheumatoid Arthritis: A Retrospective Single-Centre Cohort Study of 44 Patients in Morocco

**DOI:** 10.64898/2026.08.27.26361508

**Authors:** Najoua Ghani

## Abstract

**Background:** Tocilizumab (TCZ), a monoclonal antibody directed against the interleukin-6 receptor, is used in rheumatoid arthritis (RA) after inadequate response or secondary loss of response to conventional synthetic and biological disease-modifying antirheumatic drugs (DMARDs). Real-world data from North African cohorts remain scarce. We assessed the effectiveness and safety of TCZ in routine care and explored baseline factors associated with 6-month outcomes.

**Methods:** We conducted a retrospective, single-centre cohort study of 44 consecutive patients with RA treated with TCZ between April 2019 and January 2024 in the Department of Rheumatology, Moulay Ismail Hospital, Meknes, Morocco. Demographic, clinical, laboratory, treatment and follow-up data were extracted from medical records using a standardised electronic form. The primary effectiveness outcome was the European Alliance of Associations for Rheumatology (EULAR) response at 6 months; DAS28-ESR remission was defined as DAS28-ESR below 2.6. Safety outcomes comprised infections, neutropenia, liver-enzyme abnormalities and lipid abnormalities. Longitudinal changes were compared with the Wilcoxon signed-rank test, and associations between baseline variables dichotomised at their median and 6-month outcomes were examined with chi-square tests, in SPSS version 29.

**Results:** The cohort comprised 33 women (75.0%), with a median age of 57 years (range 32– 82) and a mean RA duration of 12.97±9.1 years. Patients had received a mean of 2.5±1.8 previous conventional DMARDs, and 41 (93.2%) had received at least one previous biological agent, including two or more tumour necrosis factor (TNF) inhibitors in 36 (81.8%). At 6 months, outcome data were available for 34 patients: 23 (67.6%) achieved a good EULAR response, 6 (17.6%) a moderate response and 5 (14.7%) no response; 12 (35.3%) were in DAS28-ESR remission. Mean DAS28-ESR fell from 5.10±1.18 at baseline to 2.74±1.38 at 6 months and 2.45±1.33 at 12 months, and the mean prednisone-equivalent dose fell from 8.3±7.1 to 5 mg/day. Twenty-two infectious episodes were recorded, including one serious infection (purulent pleurisy) requiring hospitalisation; 5 patients (11.4%) had a temporary interruption and 1 (2.3%) a permanent discontinuation for hepatic cytolysis. A neutrophil count below 1,500/mm³ occurred in 13 patients (29.5%), with no count below 1,000/mm³, while mean neutrophils declined from 6.3±3.0 to 2.6±1.2 G/L at 12 months. Mean LDL cholesterol rose from 1.18 to 1.49 g/L and HDL cholesterol from 0.58 to 0.82 g/L. Rheumatoid-factor positivity was the only baseline variable associated with the EULAR response category (p=0.007); a baseline tender joint count above six was associated with a lower remission rate (23.5%, p=0.007), as was, borderline, a pain visual analogue scale above 65 mm (31.2%, p=0.05).

**Conclusions:** In this heavily pretreated real-world RA cohort, TCZ was associated with a substantial and sustained reduction in disease activity and a manageable safety profile consistent with its known signals. A high baseline articular and pain burden was associated with a lower probability of remission. The small sample, incomplete 6-month follow-up, retrospective design and absence of adjusted effect estimates limit interpretation, and the reported associations should be regarded as hypothesis-generating.

## 1. Introduction

Rheumatoid arthritis (RA) is a chronic systemic inflammatory disease characterised by persistent synovitis, progressive structural joint damage, functional disability, impaired quality of life and excess cardiovascular morbidity. Contemporary management follows a treat-to-target strategy combining conventional synthetic, biological and targeted synthetic disease-modifying antirheumatic drugs (DMARDs), with treatment escalated until sustained remission or low disease activity is achieved [1–3].

Despite this strategy, a substantial proportion of patients experience primary non-response or secondary loss of response to tumour necrosis factor (TNF) inhibitors. This has driven the use of agents directed at alternative inflammatory pathways. Interleukin-6 (IL-6) is a pleiotropic cytokine central to acute and chronic inflammation, immune-cell differentiation, the hepatic acute-phase response, synovial inflammation and the systemic manifestations of RA, including anaemia, fatigue and osteoporosis [4,5]. Tocilizumab (TCZ), a humanised monoclonal antibody against the IL-6 receptor, is an established biological DMARD for patients with an inadequate response to conventional synthetic or other biological therapies, and is one of the few biologics with demonstrated superiority over anti-TNF monotherapy [2,3,6–8].

Pivotal randomised trials were conducted in selected populations under controlled conditions, and their findings are not automatically transferable to routine care, where patients are older, more comorbid, more heavily pretreated and less closely monitored. Real-world data from North African settings, where access to biological therapy, monitoring capacity and treatment sequencing differ from those in European and North American registries, remain particularly scarce.

The objectives of this study were to evaluate the real-world effectiveness and safety of TCZ in a Moroccan cohort of patients with refractory RA over 12 months of follow-up, and to explore baseline factors associated with the EULAR response and with DAS28-ESR remission at 6 months.

## 2. Methods

### 2.1 Study design and setting

This was a retrospective, single-centre observational cohort study conducted in the Department of Rheumatology of Moulay Ismail Hospital, Meknes, Morocco, a referral centre for inflammatory rheumatic disease in the Fes–Meknes region. The study is reported in accordance with the STROBE (Strengthening the Reporting of Observational Studies in Epidemiology) recommendations for cohort studies.

### 2.2 Participants

All consecutive adult patients with a physician-confirmed diagnosis of RA who received at least one infusion of TCZ between April 2019 and January 2024 were eligible. TCZ was prescribed in routine care after an inadequate response or a secondary loss of response to conventional synthetic DMARDs and, in most patients, to one or more biological agents. No restriction was applied with respect to disease duration, number of previous DMARDs, serological status or TCZ dose. Forty-four patients met these criteria and constituted the analysis cohort. No patient was excluded after inclusion.

### 2.3 Data sources and measurement

Data were extracted from electronic medical records and, where necessary, from archived paper source records, using a standardised electronic data-collection form completed by the investigator. Collected variables comprised demographic characteristics (sex, age, weight), disease characteristics (RA duration, rheumatoid factor and anti-citrullinated protein antibody status, erosive disease), previous and concomitant treatment (conventional synthetic DMARDs, biological agents, corticosteroid dose expressed as prednisone equivalent), and clinical and laboratory measures recorded at baseline and at months 1, 3, 6, 9 and 12.

### 2.4 Treatment and follow-up

TCZ was administered intravenously every four weeks at 8 mg/kg in 43 patients and at 4 mg/kg in one patient, in accordance with the prescribing physician’s judgement and the summary of product characteristics. Concomitant conventional synthetic DMARD therapy and corticosteroid use were continued or adjusted at the discretion of the treating physician. Complete blood counts, liver-enzyme measurements and lipid profiles were obtained before each infusion in accordance with routine monitoring practice, allowing detection of laboratory abnormalities from the first month of exposure onwards.

### 2.5 Outcomes

The primary effectiveness outcome was the EULAR response at 6 months, categorised as good, moderate or no response according to the magnitude of change in DAS28-ESR and the DAS28-ESR value attained. The secondary effectiveness outcome was DAS28-ESR remission, defined as DAS28-ESR below 2.6. Additional descriptive outcomes comprised pain on a 0–100 mm visual analogue scale (VAS), tender joint count (TJC, 28 joints), swollen joint count (SJC, 28 joints), erythrocyte sedimentation rate (ESR), C-reactive protein (CRP), DAS28-ESR, neutrophil and platelet counts, aspartate and alanine aminotransferase (AST, ALT), the lipid profile, and daily prednisone-equivalent corticosteroid dose.

Safety outcomes comprised all infectious episodes recorded during follow-up (classified as serious when they required hospitalisation, intravenous antimicrobial therapy or resulted in death), neutropenia (defined by thresholds of 1,500/mm³ and 1,000/mm³), elevation of transaminases above the laboratory upper limit of normal (ULN), lipid abnormalities, and any TCZ dose reduction, temporary interruption or permanent discontinuation attributable to an adverse event.

### 2.6 Handling of missing data

All available observations were analysed as recorded, without imputation. Denominators therefore vary across time points and are reported explicitly for every outcome. At 6 months, complete effectiveness data were available for 34 of the 44 patients (77.3%); the remaining 10 patients had either discontinued treatment, were lost to follow-up, or had not yet reached the 6-month time point at the census date. Because the number of patients contributing laboratory values also fell over time, baseline summary statistics computed on the whole cohort (Table 1) differ slightly from the baseline values of the longitudinal series (Table 3), which are restricted to patients with repeated measurements. Analyses of 6-month outcomes are restricted to patients with observed data, an approach that assumes data are missing at random and that may overestimate effectiveness if patients with a poor response were preferentially lost to follow- up.

**Table 1.**
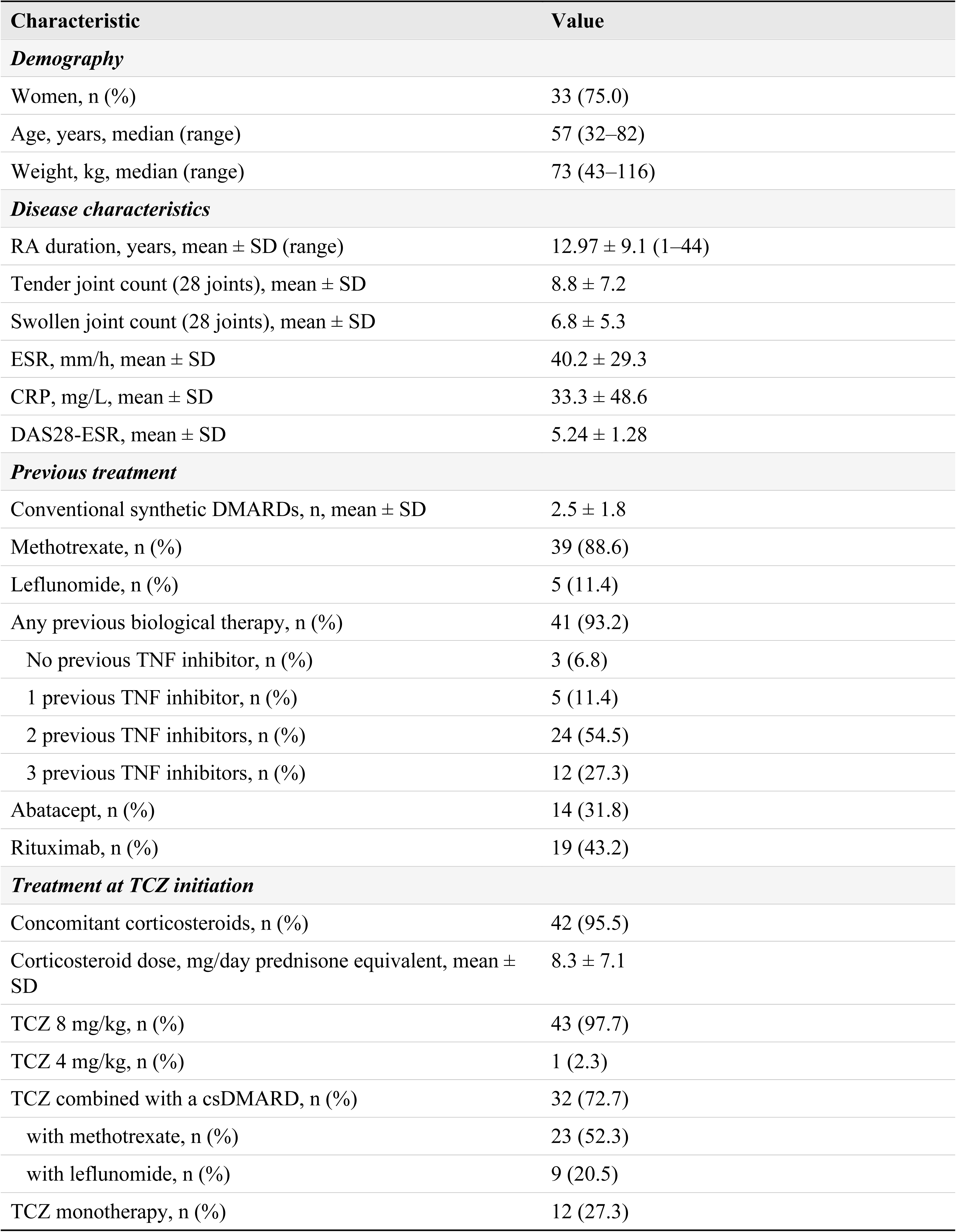
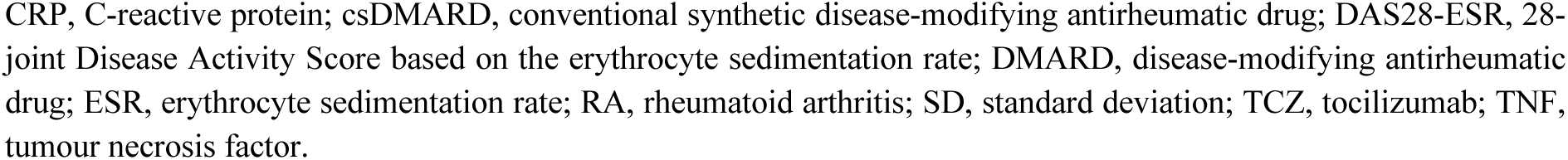
Baseline demographic, clinical and treatment characteristics of the cohort (n=44)

| Characteristic | Value |
| --- | --- |
| **Demography** |  |
| Women, n (%) | 33 (75.0) |
| Age, years, median (range) | 57 (32–82) |
| Weight, kg, median (range) | 73 (43–116) |
| **Disease characteristics** |  |
| RA duration, years, mean ± SD (range) | $12.97 \pm 9.1$ (1–44) |
| Tender joint count (28 joints), mean ± SD | $8.8 \pm 7.2$ |
| Swollen joint count (28 joints), mean ± SD | $6.8 \pm 5.3$ |
| ESR, mm/h, mean ± SD | $40.2 \pm 29.3$ |
| CRP, mg/L, mean ± SD | $33.3 \pm 48.6$ |
| DAS28-ESR, mean ± SD | $5.24 \pm 1.28$ |
| **Previous treatment** |  |
| Conventional synthetic DMARDs, n, mean ± SD | $2.5 \pm 1.8$ |
| Methotrexate, n (%) | 39 (88.6) |
| Leflunomide, n (%) | 5 (11.4) |
| Any previous biological therapy, n (%) | 41 (93.2) |
| No previous TNF inhibitor, n (%) | 3 (6.8) |
| 1 previous TNF inhibitor, n (%) | 5 (11.4) |
| 2 previous TNF inhibitors, n (%) | 24 (54.5) |
| 3 previous TNF inhibitors, n (%) | 12 (27.3) |
| Abatacept, n (%) | 14 (31.8) |
| Rituximab, n (%) | 19 (43.2) |
| **Treatment at TCZ initiation** |  |
| Concomitant corticosteroids, n (%) | 42 (95.5) |
| Corticosteroid dose, mg/day prednisone equivalent, mean ± SD | $8.3 \pm 7.1$ |
| TCZ 8 mg/kg, n (%) | 43 (97.7) |
| TCZ 4 mg/kg, n (%) | 1 (2.3) |
| TCZ combined with a csDMARD, n (%) | 32 (72.7) |
| with methotrexate, n (%) | 23 (52.3) |
| with leflunomide, n (%) | 9 (20.5) |
| TCZ monotherapy, n (%) | 12 (27.3) |

CRP, C-reactive protein; csDMARD, conventional synthetic disease-modifying antirheumatic drug; DAS28-ESR, 28-joint Disease Activity Score based on the erythrocyte sedimentation rate; DMARD, disease-modifying antirheumatic drug; ESR, erythrocyte sedimentation rate; RA, rheumatoid arthritis; SD, standard deviation; TCZ, tocilizumab; TNF, tumour necrosis factor.

### 2.7 Statistical analysis

Analyses were performed with IBM SPSS Statistics version 29 (IBM Corp., Armonk, NY, USA). Continuous variables are summarised as mean ± standard deviation, or as median with range where the distribution was skewed; categorical variables are summarised as counts and percentages, the denominator being stated in each case. Longitudinal changes in clinical and laboratory measures were compared with the Wilcoxon signed-rank test for paired data. Associations between baseline variables and 6-month outcomes were examined using Pearson chi-square tests, with continuous predictors dichotomised at their observed median values (TJC >6, SJC >6, ESR >34 mm/h, CRP >12 mg/L, DAS28-ESR >5.3, pain VAS >65 mm). A two-sided p value below 0.05 was considered to indicate statistical significance.

Given the sample size and the number of comparisons performed, no multivariable model was fitted and no adjusted effect estimates, odds ratios or confidence intervals were computed. No correction for multiple testing was applied. All association analyses are therefore exploratory and hypothesis-generating rather than confirmatory, and the reported p values should be interpreted descriptively.

### 2.8 Ethics and informed consent

This study consisted of a retrospective analysis of data generated during routine clinical care. It was non-interventional: no procedure, investigation or treatment was performed for research purposes, and patient management was at no point modified as a result of the study. All records were irreversibly anonymised before analysis, and no directly or indirectly identifying information — name, initials, medical record number, date of birth, exact age or address — was retained in the analysis dataset.

The study protocol was reviewed by the Research Ethics Committee of the Faculty of Medicine, Euromed University of Fez, Morocco, which granted an exemption from full ethics review and waived the requirement for individual written informed consent, the irreversibly anonymised nature of the analysis dataset precluding re-identification of, and any subsequent contact with, the patients concerned. The study was conducted in accordance with the principles of the Declaration of Helsinki. No information permitting the identification of an individual patient is reported.

## 3. Results

### 3.1 Cohort characteristics

Forty-four patients were included, of whom 33 (75.0%) were women. The median age was 57 years (range 32–82) and the median weight 73 kg (range 43–116). The mean duration of RA at TCZ initiation was 12.97±9.1 years (range 1–44), indicating a long-standing and established disease population.

Patients were heavily pretreated: they had received a mean of 2.5±1.8 previous conventional synthetic DMARDs, and 41 (93.2%) had received at least one previous biological agent. Concomitant corticosteroids were used by 42 patients (95.5%) at a mean prednisone-equivalent dose of 8.3±7.1 mg/day. Baseline disease activity was high, with a mean TJC of 8.8±7.2, a mean SJC of 6.8±5.3, a mean ESR of 40.2±29.3 mm/h, a mean CRP of 33.3±48.6 mg/L and a mean DAS28-ESR of 5.24±1.28. Baseline characteristics are detailed in Table 1.

### 3.2 Previous and concomitant treatment

Thirty-nine patients (88.6%) had previously received methotrexate and five (11.4%) leflunomide. Most patients had been exposed to more than one TNF inhibitor before TCZ: three patients (6.8%) had received none, 5 (11.4%) one agent, 24 (54.5%) two agents and 12 (27.3%) three agents, so that 36 patients (81.8%) had failed at least two TNF inhibitors. Previous exposure to abatacept and rituximab was recorded in 14 (31.8%) and 19 (43.2%) patients, respectively.

TCZ was combined with a conventional synthetic DMARD in 32 patients (72.7%), namely methotrexate in 23 (52.3%) and leflunomide in 9 (20.5%). Twelve patients (27.3%) received TCZ as monotherapy, most often because of previous methotrexate intolerance.

Treatment was discontinued in six patients during follow-up: in three patients at month 8 following sustained remission, in two because of inefficacy (after the seventh and ninth infusions, respectively), and in one because of hepatic intolerance (after the tenth infusion).

### 3.3 Effectiveness

At month 3, EULAR response data were available for 42 patients: 25 (59.5%) had a good response, 14 (33.3%) a moderate response and 3 (7.1%) no response. At month 6, data were available for 34 patients: 23 (67.6%) had a good response, 6 (17.6%) a moderate response and 5 (14.7%) no response. Twelve of these 34 patients (35.3%) were in DAS28-ESR remission at month 6. The distribution of EULAR response categories at both time points is shown in Figure 1.

**Figure 1.**
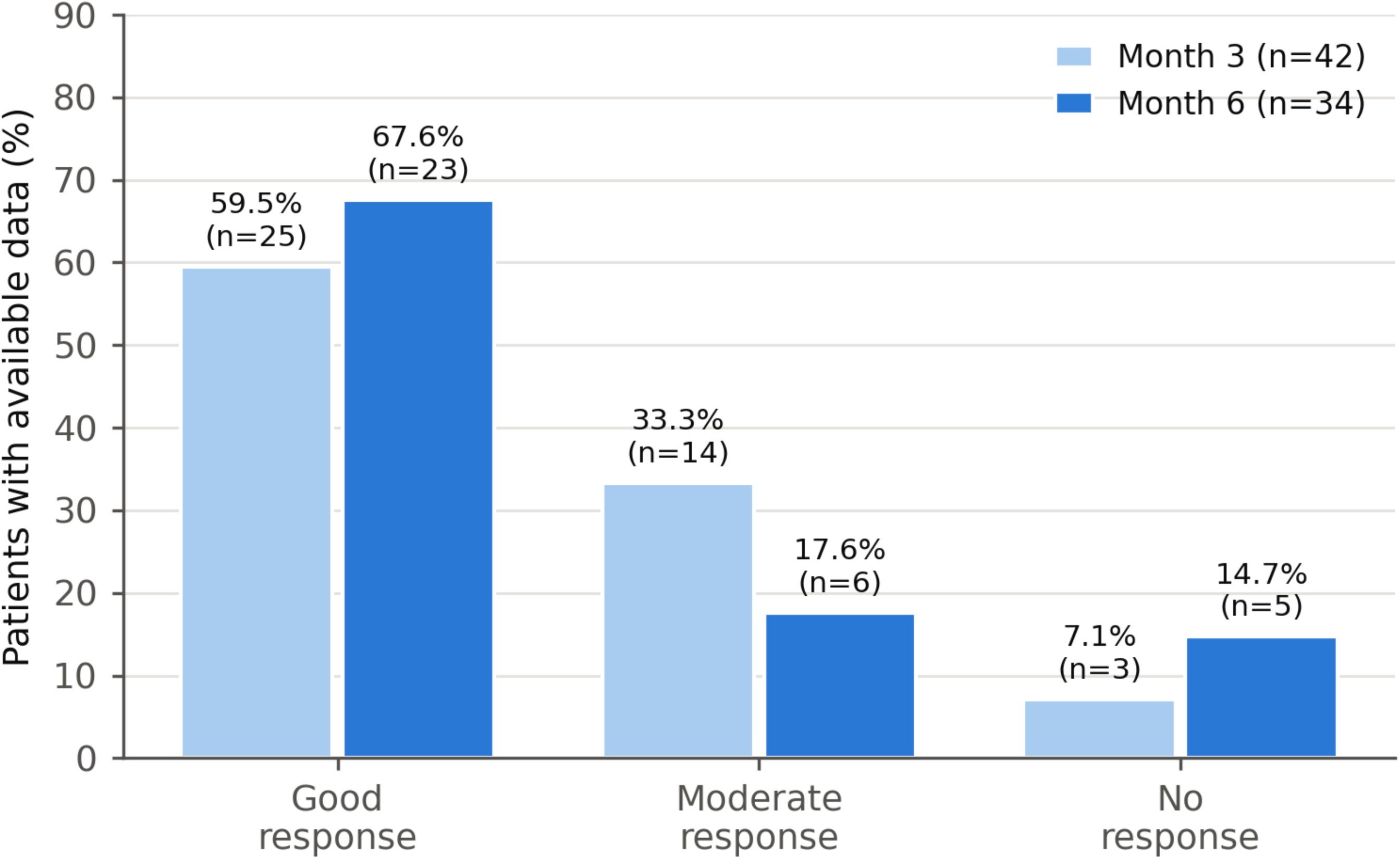
Distribution of EULAR response categories at month 3 (n=42) and month 6 (n=34). Bars show the proportion of patients with available data in each response category; counts are given in parentheses. EULAR, European Alliance of Associations for Rheumatology.

Disease activity decreased progressively throughout follow-up (Table 3, Figure 2). Mean DAS28-ESR fell from 5.10±1.18 at baseline among patients with longitudinal data to 4.10±1.52 at month 1, 3.33±1.38 at month 3, 2.74±1.38 at month 6, 3.12±1.60 at month 9 and 2.45±1.33 at month 12. Mean pain VAS decreased from 67 mm at baseline to 30 mm at month 3. The steepest fall in acute-phase reactants occurred during the first month, with mean ESR declining from 39.7±28.6 to 15.6±16.6 mm/h and mean CRP from 23.6±25.0 to 13.1±39.8 mg/L; both continued to decline thereafter, reaching 6.9±7.1 mm/h and 3.1±2.7 mg/L at month 12. Paired comparisons across time points were significant at p<0.05 (Wilcoxon signed-rank test).

**Figure 2.**
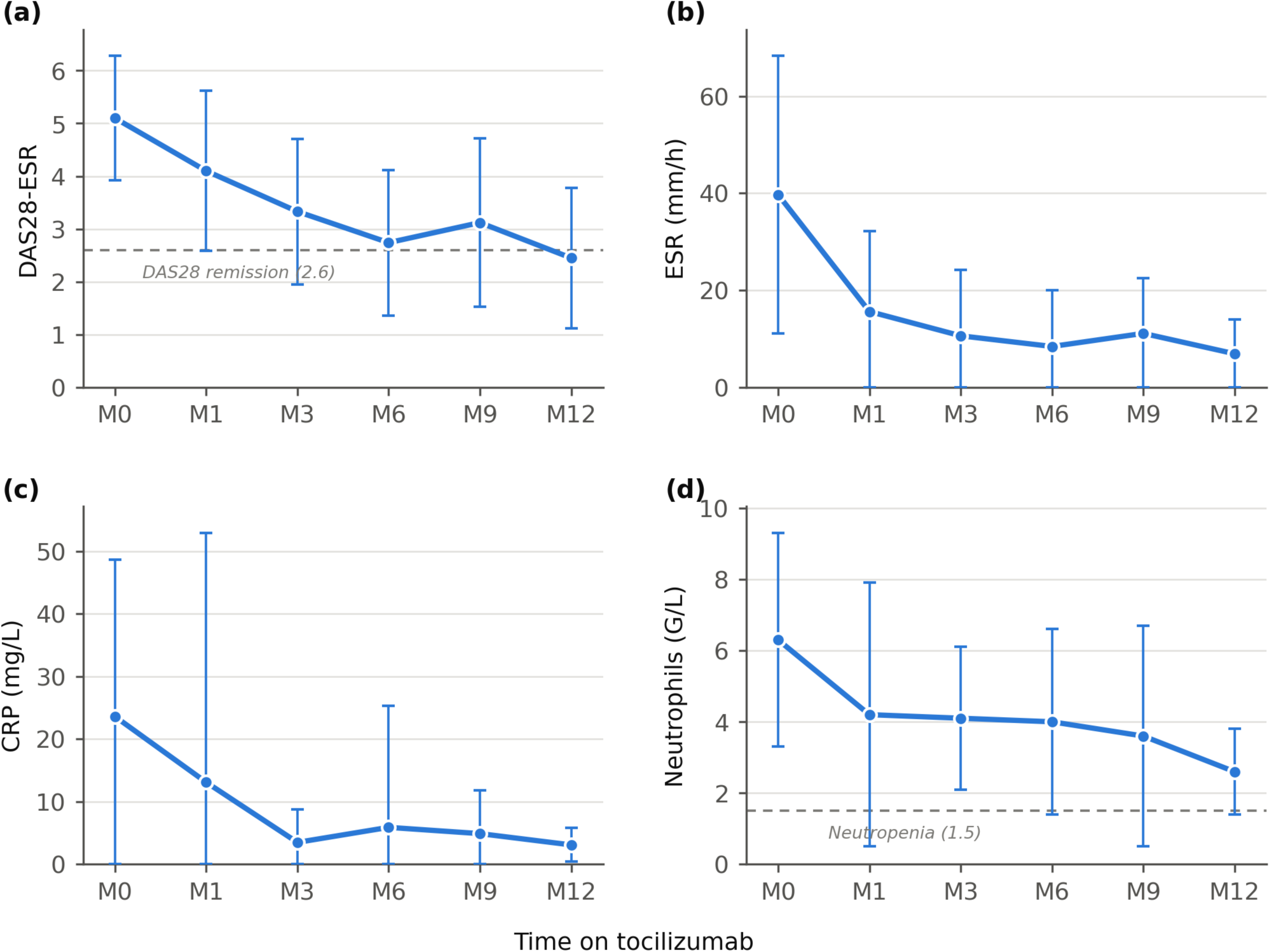
Longitudinal changes from baseline to month 12 in (a) DAS28-ESR, (b) erythrocyte sedimentation rate, (c) C-reactive protein and (d) neutrophil count during tocilizumab treatment. Points are means and bars are standard deviations among patients with an available measurement at each time point; the lower bound of each bar is truncated at zero where the standard deviation exceeds the mean. Dashed lines mark the DAS28-ESR remission threshold (2.6) and the neutropenia threshold (1.5 G/L). Paired comparisons across time points were significant at p<0.05 (Wilcoxon signed-rank test). CRP, C-reactive protein; DAS28-ESR, 28-joint Disease Activity Score based on the erythrocyte sedimentation rate; ESR, erythrocyte sedimentation rate.

The mean daily prednisone-equivalent corticosteroid dose fell from 8.3±7.1 mg/day at baseline to 5 mg/day at month 6, indicating a corticosteroid-sparing effect. Six-month effectiveness outcomes are summarised in Table 2.

**Table 2.** Effectiveness outcomes during tocilizumab treatment.

| Outcome | Result | Patients with available data |
| --- | --- | --- |
| EULAR response, month 3 | Good 25 (59.5%); moderate 14 (33.3%); none 3 (7.1%) | n=42 |
| EULAR response, month 6 | Good 23 (67.6%); moderate 6 (17.6%); none 5 (14.7%) | n=34 |
| DAS28-ESR remission ($< 2.6$), month 6 | 12 (35.3%) | n=34 |
| DAS28-ESR, mean ± SD | Baseline $5.10 \pm 1.18$; month 6, $2.74 \pm 1.38$; month 12, $2.45 \pm 1.33$ | Patients with paired data at each time point |
| Pain VAS (0–100 mm), mean | Baseline 67; month 3, 30 | Dispersion not recorded |
| Corticosteroid dose, mg/day | Baseline $8.3 \pm 7.1$; month 6, 5.0 | Prednisone equivalent |
| Discontinuation during follow-up | 6 patients: remission 3; inefficacy 2; hepatic intolerance 1 | n=44 |

DAS28-ESR, 28-joint Disease Activity Score based on the erythrocyte sedimentation rate; EULAR, European Alliance of Associations for Rheumatology; SD, standard deviation; VAS, visual analogue scale.

### 3.4 Safety

Twenty-two infectious or mucocutaneous episodes were recorded during follow-up: bronchitis in 10, gastroenteritis in 4, cutaneous or urticarial reactions in 3, ear-nose-throat infection (sinusitis, otitis) in 2, purulent pleurisy in 1, conjunctivitis in 1 and herpetic keratitis in 1.

A single serious adverse event requiring hospitalisation occurred. A woman in her late forties with seropositive erosive RA developed purulent pleurisy after her second TCZ infusion. No causative organism was identified. She received amoxicillin–clavulanic acid for three weeks, including a two-week hospital stay, recovered fully, and subsequently resumed TCZ at the same dose without recurrence.

Adverse events led to a TCZ dose reduction in 4 patients (9.1%), comprising three episodes of neutropenia and one prostatitis, and to a temporary interruption of treatment in 5 patients (11.4%), comprising worsening of an associated condition in three, a non-healing wound in one and herpetic keratitis in one. One patient (2.3%) discontinued TCZ permanently because of hepatic cytolysis with ALT reaching four times the ULN.

At least one neutrophil count below 1,500/mm³ was recorded in 13 patients (29.5%); no patient had a count below 1,000/mm³. These episodes were transient and were not associated with a clinically significant infection. Across the cohort, the mean neutrophil count declined progressively from 6.3±3.0 G/L at baseline to 4.2±3.7 G/L at month 1 and 2.6±1.2 G/L at month 12, and the mean platelet count from 328.5±105 to 198.0±22.5 G/L, both remaining within or close to the normal range (Table 3, Figure 2d).

**Table 3.** Longitudinal clinical and laboratory parameters from baseline to month 12.

| Time point | DAS28-ESR | ESR (mm/h) | CRP (mg/L) | Neutrophils (G/L) | Platelets (G/L) | ALT (IU/L) |
| --- | --- | --- | --- | --- | --- | --- |
| Baseline | $5.10 \pm 1.18$ | $39.7 \pm 28.6$ | $23.6 \pm 25.0$ | $6.3 \pm 3.0$ | $328.5 \pm 105$ | $22.6 \pm 9.7$ |
| Month 1 | $4.10 \pm 1.52$ | $15.6 \pm 16.6$ | $13.1 \pm 39.8$ | $4.2 \pm 3.7$ | $245.3 \pm 73.3$ | $28.4 \pm 12.8$ |
| Month 3 | $3.33 \pm 1.38$ | $10.6 \pm 13.6$ | $3.5 \pm 5.3$ | $4.1 \pm 2.0$ | $250.5 \pm 53.8$ | $28.4 \pm 12.3$ |
| Month 6 | $2.74 \pm 1.38$ | $8.4 \pm 11.6$ | $5.9 \pm 19.4$ | $4.0 \pm 2.6$ | $241.3 \pm 66.8$ | $29.0 \pm 15.2$ |
| Month 9 | $3.12 \pm 1.60$ | $11.1 \pm 11.4$ | $4.9 \pm 6.9$ | $3.6 \pm 3.1$ | $257.8 \pm 79.5$ | $36.9 \pm 26.9$ |
| Month 12 | $2.45 \pm 1.33$ | $6.9 \pm 7.1$ | $3.1 \pm 2.7$ | $2.6 \pm 1.2$ | $198.0 \pm 22.5$ | $32.5 \pm 12.5$ |

Values are mean ± standard deviation among patients with an available measurement at each time point. Mean aspartate aminotransferase was $20.8 \pm 6.2$ IU/L at baseline, $23.1 \pm 8.3$ at month 1, $23.8 \pm 6.0$ at month 3, $24.6 \pm 8.0$ at month 6, $28.1 \pm 13.4$ at month 9 and $26.5 \pm 9.1$ IU/L at month 12. ALT, alanine aminotransferase; CRP, C-reactive protein; DAS28-ESR, 28-joint Disease Activity Score based on the erythrocyte sedimentation rate; ESR, erythrocyte sedimentation rate.

Transaminase elevation above the ULN but below twice the ULN was observed in 7 patients (15.9%) and resolved without intervention in all but one. Mean ALT rose modestly from 22.6±9.7 IU/L at baseline to a peak of 36.9±26.9 IU/L at month 9. One patient developed a progressive rise in ALT, reaching three times and subsequently four times the ULN by month 9. Methotrexate was withdrawn and TCZ stopped; a liver biopsy demonstrated steatosis considered probably metabolic in origin, and liver-function tests normalised after both drugs were discontinued.

Mean LDL cholesterol increased from 1.18±0.60 g/L at baseline to 1.49±0.69 g/L at month 12, and mean HDL cholesterol from 0.58 to 0.82 g/L, while mean triglycerides fluctuated between 1.12 and 1.73 g/L (Table 5, Figure 3). The number of patients with an LDL cholesterol concentration of 1.6 g/L or above was small at every time point, peaking at six patients at months 3 and 6. Twelve patients (27.3%) received lipid-lowering therapy, of whom eight started treatment between months 2 and 6 and four were already receiving a statin before TCZ initiation. No cardiovascular event was recorded during follow-up. Safety findings are summarised in Table 4.

**Figure 3.**
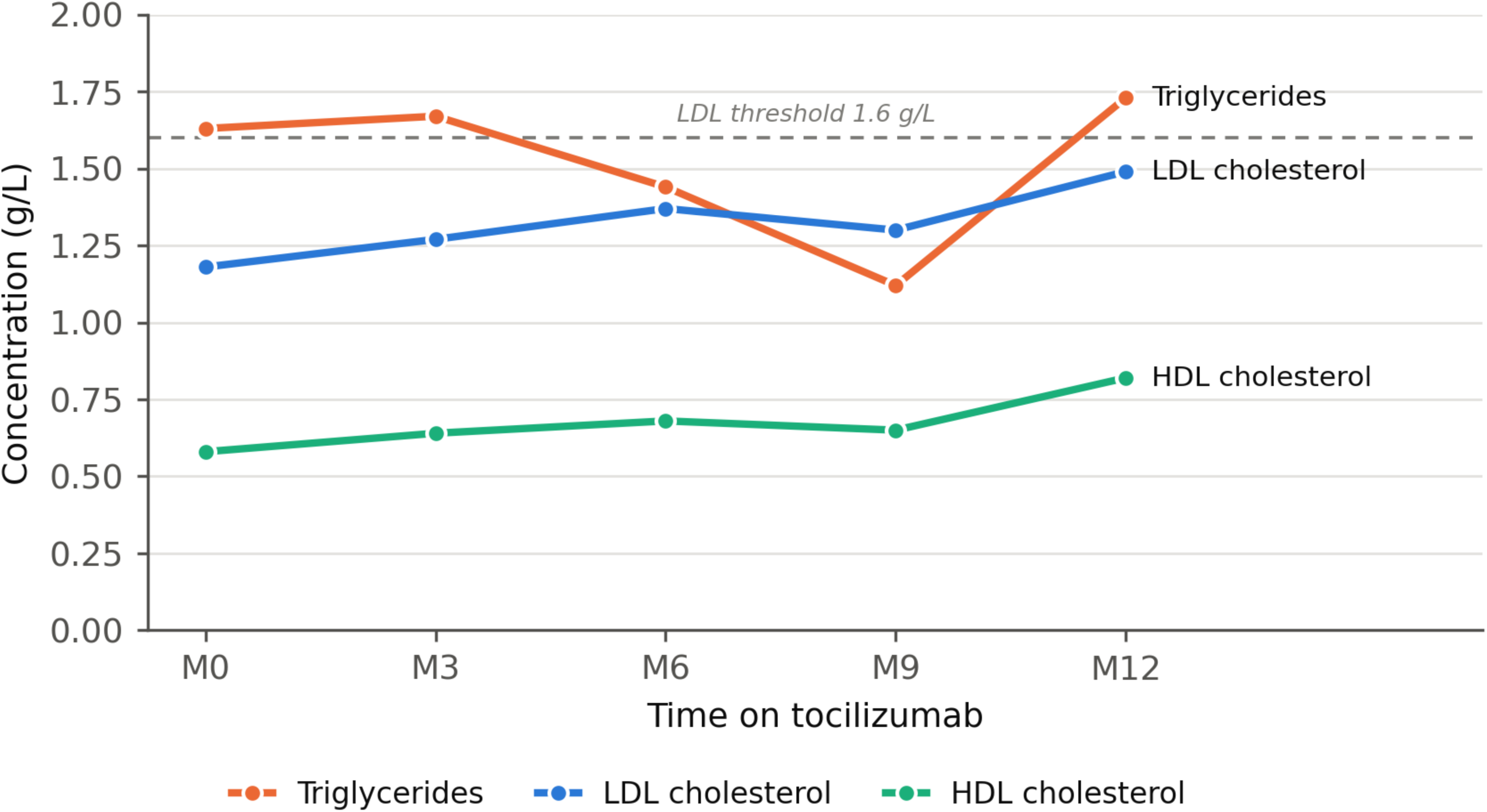
Longitudinal changes in the mean lipid profile during tocilizumab treatment. Values are means among patients with an available measurement at each time point. The dashed line marks the 1.6 g/L LDL cholesterol threshold used to classify elevated values in Table 5. HDL, high-density lipoprotein; LDL, low- density lipoprotein.

**Table 4.** Adverse events and infectious episodes during tocilizumab treatment (n=44)

| Event | n (%) | Detail |
| --- | --- | --- |
| *Management of adverse events* |  |  |
| Dose reduction | 4 (9.1) | Neutropenia 3; prostatitis 1 |
| Temporary interruption | 5 (11.4) | Worsening of an associated condition 3; non-healing wound 1; herpetic keratitis 1 |
| Permanent discontinuation | 1 (2.3) | Hepatic cytolysis (ALT 4× ULN) |
| *Infectious and mucocutaneous episodes (22 in total)* |  |  |
| Bronchitis | 10 | — |
| Gastroenteritis | 4 | — |
| Cutaneous / urticarial | 3 | Non-infectious mucocutaneous reactions |
| Ear-nose-throat (sinusitis, otitis) | 2 | — |
| Purulent pleurisy | 1 | Serious; hospitalisation; no organism identified; TCZ resumed |
| Conjunctivitis | 1 | — |
| Herpetic keratitis | 1 | Led to temporary interruption |
| *Laboratory abnormalities* |  |  |
| Neutrophils <1,500/mm<sup>3</sup> | 13 (29.5) | Transient; no associated clinical infection |
| Neutrophils <1,000/mm<sup>3</sup> | 0 | — |
| Transaminases >ULN and <2× ULN | 7 (15.9) | Self-limiting in all but one patient |
| ALT ≥3× ULN | 1 (2.3) | Biopsy-proven steatosis, probably metabolic |
| Lipid-lowering therapy | 12 (27.3) | 8 initiated between months 2 and 6; 4 pre-existing |

ALT, alanine aminotransferase; TCZ, tocilizumab; ULN, upper limit of normal.

**Table 5.**
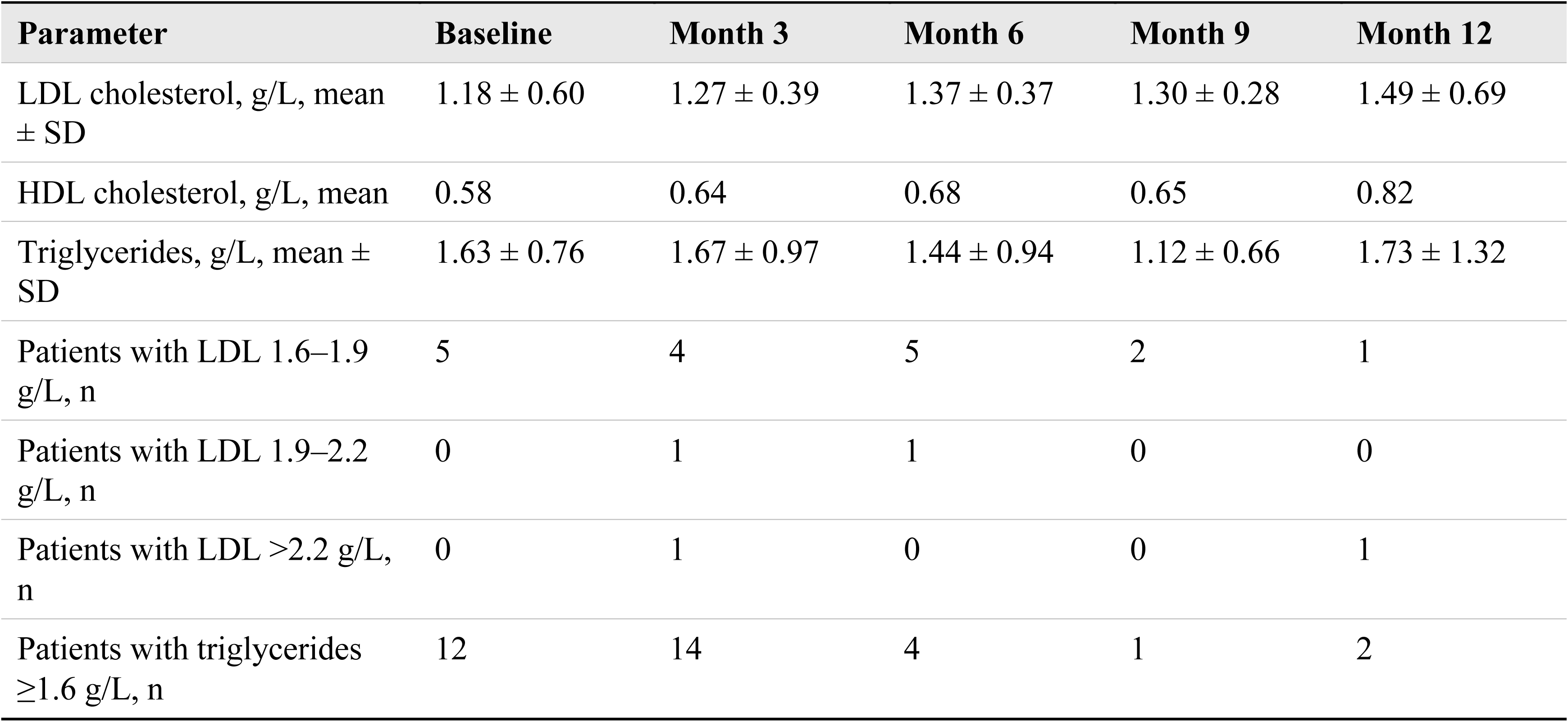
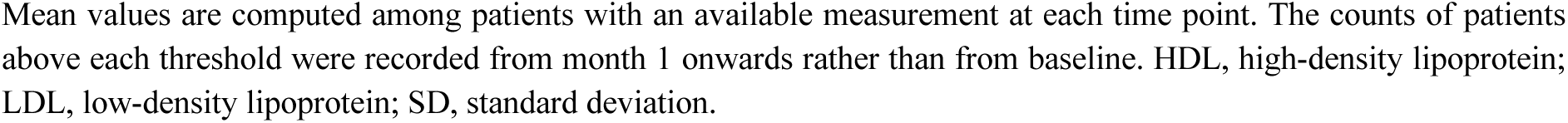
Lipid profile from baseline to month 12.

| Parameter | Baseline | Month 3 | Month 6 | Month 9 | Month 12 |
| --- | --- | --- | --- | --- | --- |
| LDL cholesterol, g/L, mean ± SD | 1.18 ± 0.60 | 1.27 ± 0.39 | 1.37 ± 0.37 | 1.30 ± 0.28 | 1.49 ± 0.69 |
| HDL cholesterol, g/L, mean | 0.58 | 0.64 | 0.68 | 0.65 | 0.82 |
| Triglycerides, g/L, mean ± SD | 1.63 ± 0.76 | 1.67 ± 0.97 | 1.44 ± 0.94 | 1.12 ± 0.66 | 1.73 ± 1.32 |
| Patients with LDL 1.6–1.9 g/L, n | 5 | 4 | 5 | 2 | 1 |
| Patients with LDL 1.9–2.2 g/L, n | 0 | 1 | 1 | 0 | 0 |
| Patients with LDL >2.2 g/L, n | 0 | 1 | 0 | 0 | 1 |
| Patients with triglycerides ≥1.6 g/L, n | 12 | 14 | 4 | 1 | 2 |

Mean values are computed among patients with an available measurement at each time point. The counts of patients above each threshold were recorded from month 1 onwards rather than from baseline. HDL, high-density lipoprotein; LDL, low-density lipoprotein; SD, standard deviation.

### 3.5 Baseline factors associated with 6-month outcomes

Rheumatoid-factor positivity was the only baseline variable associated with the EULAR response category at 6 months: among rheumatoid-factor-positive patients, 72% were good responders, 16% moderate responders and 12% non-responders (p=0.007). None of the other variables examined was associated with the response category, including a tender joint count above six (p=0.50), a swollen joint count above six (p=0.28), an ESR above 34 mm/h (p=0.30), a CRP above 12 mg/L (p=0.53), a DAS28-ESR above 5.3 (p=0.63) and concomitant methotrexate (p=0.50), although patients receiving concomitant methotrexate had the highest observed proportion of good responders (84%). These results are presented in Table 6.

**Table 6.**
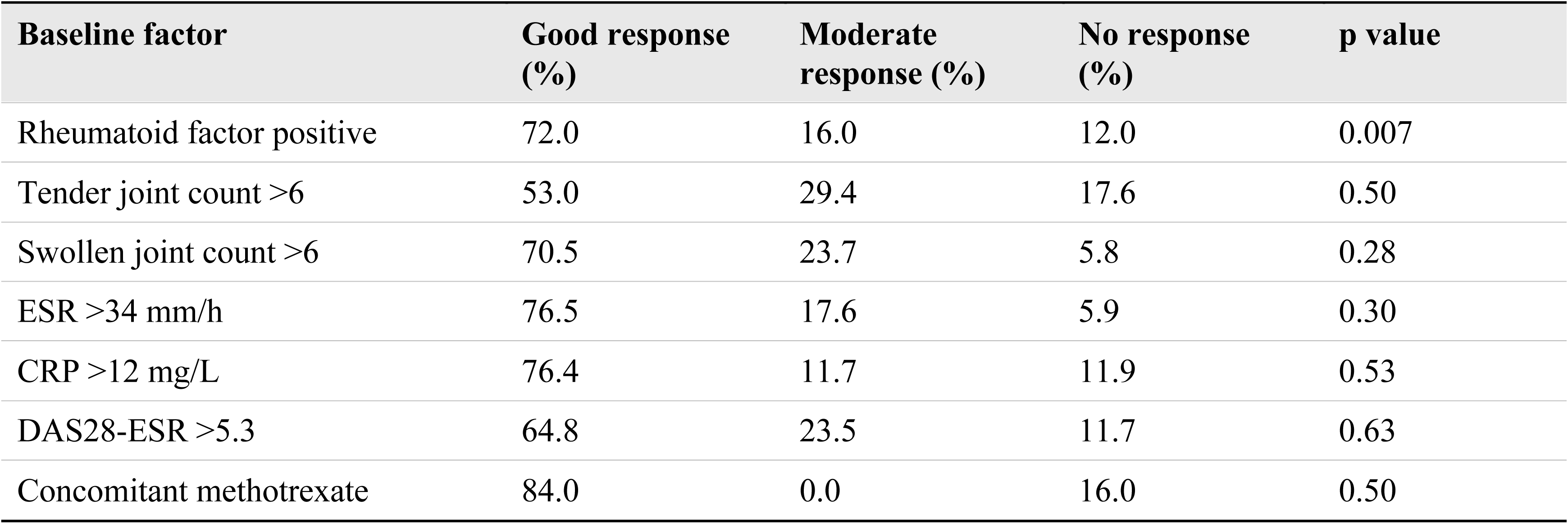
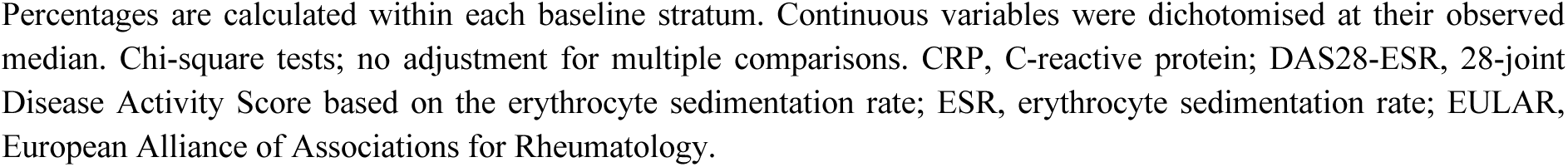
Baseline factors and EULAR response category at 6 months (bivariate chi-square analyses, n=34)

| Baseline factor | Good response (%) | Moderate response (%) | No response (%) | p value |
| --- | --- | --- | --- | --- |
| Rheumatoid factor positive | 72.0 | 16.0 | 12.0 | 0.007 |
| Tender joint count >6 | 53.0 | 29.4 | 17.6 | 0.50 |
| Swollen joint count >6 | 70.5 | 23.7 | 5.8 | 0.28 |
| ESR >34 mm/h | 76.5 | 17.6 | 5.9 | 0.30 |
| CRP >12 mg/L | 76.4 | 11.7 | 11.9 | 0.53 |
| DAS28-ESR >5.3 | 64.8 | 23.5 | 11.7 | 0.63 |
| Concomitant methotrexate | 84.0 | 0.0 | 16.0 | 0.50 |

Percentages are calculated within each baseline stratum. Continuous variables were dichotomised at their observed median. Chi-square tests; no adjustment for multiple comparisons. CRP, C-reactive protein; DAS28-ESR, 28-joint Disease Activity Score based on the erythrocyte sedimentation rate; ESR, erythrocyte sedimentation rate; EULAR, European Alliance of Associations for Rheumatology.

The pattern differed for DAS28-ESR remission at 6 months (Table 7). A baseline tender joint count above six was associated with a markedly lower remission rate (23.5% versus 35.3% in the cohort as a whole; p=0.007), and a baseline pain VAS above 65 mm showed a borderline association in the same direction (31.2%; p=0.05). A swollen joint count above six (41.1%; p=0.36), an ESR above 34 mm/h (52.9%; p=0.36) and a CRP above 12 mg/L (47.0%; p=0.63) were not associated with remission. Thus, a high baseline articular and pain burden was associated with a lower probability of reaching the absolute remission threshold, without being associated with the magnitude of improvement captured by the EULAR response categories.

**Table 7.** Baseline factors and DAS28-ESR remission at 6 months (bivariate chi-square analyses, n=34)

| Baseline factor | Patients in remission (%) | p value |
| --- | --- | --- |
| Tender joint count >6 | 23.5 | 0.007 |
| Pain VAS >65 mm | 31.2 | 0.05 |
| Swollen joint count >6 | 41.1 | 0.36 |
| CRP >12 mg/L | 47.0 | 0.63 |
| ESR >34 mm/h | 52.9 | 0.36 |

Remission was defined as DAS28-ESR below 2.6. Percentages are calculated within each baseline stratum and should be compared with the overall 6-month remission rate of 35.3%. Continuous variables were dichotomised at their observed median. Chi-square tests; no adjustment for multiple comparisons. CRP, C-reactive protein; DAS28-ESR, 28-joint Disease Activity Score based on the erythrocyte sedimentation rate; ESR, erythrocyte sedimentation rate; VAS, visual analogue scale.

Because no adjusted effect estimates, odds ratios or confidence intervals could be computed in a sample of this size, and because no correction for multiple testing was applied, these findings are exploratory and must not be interpreted as independent predictors of response.

## 4. Discussion

In this retrospective real-world cohort of heavily pretreated patients with RA, TCZ was associated with a high rate of good EULAR response and with consistent improvement across clinical, functional and laboratory measures of disease activity. At 6 months, 67.6% of patients with available data were good EULAR responders and 35.3% were in DAS28-ESR remission, in a population in which 93.2% had already failed at least one biological agent and 81.8% had been exposed to two or more TNF inhibitors.

### 4.1 Effectiveness in context

The pivotal OPTION and RADIATE trials established the efficacy of TCZ in combination with methotrexate in patients with an inadequate response to conventional DMARDs and to TNF inhibitors, respectively, and the ADACTA trial demonstrated superiority of TCZ monotherapy over adalimumab monotherapy; a subsequent systematic review and meta-analysis confirmed these findings across trial populations [6–9]. Contemporary observational studies report sustained improvements in disease activity in routine practice, with the ARATA study documenting effectiveness maintained over two years and the TReasure registry reporting comparable outcomes when TCZ is used as a first-line biological agent [10,11]. The response rates observed here are broadly consistent with those cohorts, which is notable given the degree of prior treatment failure in our population.

Cross-study comparisons should nevertheless be interpreted with caution. Populations, background treatment, duration of follow-up, outcome definitions and the handling of missing data differ substantially between real-world cohorts, and response rates calculated among patients with observed data, as in the present study, are systematically higher than those calculated on an intention-to-treat basis with non-responder imputation.

### 4.2 Biological plausibility

The early and pronounced reductions in CRP and ESR are pharmacologically expected, since IL-6 is the principal driver of hepatic acute-phase protein synthesis and its blockade normalises these markers rapidly and independently of any change in synovial inflammation [4,5]. This effect complicates the interpretation of composite indices such as DAS28-ESR, in which the acute-phase reactant contributes directly to the score, and may inflate apparent response rates relative to those obtained with indices that exclude acute-phase reactants, such as the Clinical Disease Activity Index. Reassuringly, the concurrent reductions in tender and swollen joint counts and in the pain VAS observed in the present cohort indicate that the improvement was not confined to laboratory parameters. Real-world evidence indicates that TCZ retains effectiveness in both biologic-naive and biologic-exposed patients, although treatment retention and response vary appreciably across cohorts [10–12].

### 4.3 Safety

The safety profile observed here was dominated by infections and by transient laboratory abnormalities, and no new safety signal emerged. A single serious infection occurred, and the patient was successfully retreated after resolution. Neutropenia, transaminase elevation and dyslipidaemia are recognised class effects of IL-6 receptor blockade and are consistently reported in clinical trials and registries [7–10]. In the present cohort these abnormalities were mild and rarely required intervention: no neutrophil count fell below 1,000/mm³, and only one patient discontinued treatment permanently, for hepatic cytolysis, in a context where concomitant methotrexate and biopsy-proven metabolic steatosis were plausible contributory factors.

The progressive decline in mean neutrophil and platelet counts over 12 months is consistent with the known haematological effects of IL-6 receptor blockade and did not translate into clinically significant neutropenia or bleeding in this cohort, but it supports continued blood-count monitoring throughout treatment rather than during the first months alone.

The lipid changes observed, a rise in LDL cholesterol from 1.18 to 1.49 g/L accompanied by a rise in HDL cholesterol from 0.58 to 0.82 g/L, merit attention given the elevated cardiovascular risk inherent to RA. The concurrent increase in HDL cholesterol and the reduction in systemic inflammation may partly offset the atherogenic implication of the LDL rise, and no cardiovascular event occurred during follow-up; the short follow-up and small sample nevertheless preclude any conclusion about cardiovascular risk. A large comparative cohort study found no increase in overall serious-infection risk with TCZ relative to TNF inhibitors, although the anatomical distribution of infections differed between the two classes [13]. The absence of exposure-adjusted incidence rates and of systematically recorded denominators in the present study precludes any comparative safety conclusion.

### 4.4 Factors associated with 6-month outcomes

Rheumatoid-factor positivity was associated with the EULAR response category in bivariate analysis, a finding directionally consistent with reports that seropositive patients respond well to IL-6 receptor blockade. In contrast, a high baseline tender joint count and, marginally, a high baseline pain VAS were associated with a lower probability of DAS28-ESR remission, without being associated with the EULAR response category. This dissociation is coherent rather than contradictory: the EULAR categories incorporate the magnitude of change from baseline, so patients starting from a high articular burden can improve substantially and still be classified as good responders, whereas remission depends on the absolute value attained and is therefore harder to reach from a high baseline. Since the tender joint count and the pain VAS both reflect subjective symptom burden, the association may also partly reflect central pain sensitisation, which is known to limit the attainment of remission independently of inflammatory activity.

It should also be emphasised that DAS28-ESR remission is not interchangeable with the more stringent ACR/EULAR Boolean or index-based definitions, and systematically classifies a larger proportion of patients as being in remission [14]. Previous observational studies have identified age, baseline CRP, cardiovascular history, disability, disease activity, corticosteroid use and prior biological exposure as possible correlates of response or drug retention, but no single factor has been validated sufficiently to guide individual treatment selection [15,16]. The unadjusted associations reported here are subject to confounding, to the instability inherent in small samples, and to inflation of the type I error rate from multiple testing, and should therefore be regarded as hypothesis-generating.

### 4.5 Strengths and limitations

The principal strength of this study is that it documents both effectiveness and safety of TCZ in a difficult-to-treat, heavily pretreated RA population managed in routine care in a North African referral centre, a setting under-represented in the published literature. Consecutive inclusion of all treated patients, with no exclusion after enrolment, limits selection bias within the treated population, and structured clinical and laboratory data were collected prospectively as part of routine monitoring at six predefined time points over 12 months.

Several limitations must be acknowledged. The retrospective single-centre design precludes any causal inference and leaves the results vulnerable to confounding by indication and to incomplete or heterogeneous documentation. The sample of 44 patients is small, yielding wide uncertainty around every estimate and very limited statistical power for the association analyses; the strata compared in Tables 6 and 7 contain fewer than 20 patients each, so the corresponding p values are unstable. Effectiveness data at 6 months were available for only 34 patients (77.3%), and the analysis of observed data alone may overestimate effectiveness if patients with a poor response were preferentially lost to follow-up. No adjusted effect estimates, confidence intervals or measures of precision were computed, and no correction was made for multiple comparisons. Adverse-event denominators were not systematically recorded, and exposure-adjusted incidence rates could not be calculated, so no comparative safety conclusion is possible. The absence of a comparator arm prevents any assessment of TCZ relative to alternative treatment strategies. Finally, radiographic progression, patient-reported functional outcomes such as the Health Assessment Questionnaire, and long-term drug retention were not assessed.

## 5. Conclusions

In this retrospective single-centre cohort of 44 heavily pretreated patients with rheumatoid arthritis, tocilizumab was associated with a marked and sustained reduction in disease activity, a good EULAR response in 67.6% of evaluable patients at 6 months, DAS28-ESR remission in 35.3%, and a corticosteroid-sparing effect maintained to 12 months. The safety profile was consistent with the known signals of IL-6 receptor blockade and was dominated by transient laboratory abnormalities, with one serious infection, four dose reductions, five temporary interruptions and one permanent discontinuation for hepatic cytolysis.

Rheumatoid-factor positivity was associated with the EULAR response category, whereas a high baseline tender joint count, and, marginally, a high baseline pain score, was associated with a lower probability of remission, suggesting that the articular and symptomatic burden at initiation influences the attainment of an absolute remission threshold more than it influences the magnitude of improvement. These exploratory bivariate findings require confirmation in adequately powered studies with adjusted effect estimates. Larger prospective registries with predefined outcomes, complete follow-up, systematic adverse-event denominators and transparent statistical reporting are needed to characterise the long-term effectiveness and safety of tocilizumab in refractory rheumatoid arthritis, particularly in North African populations.

## Declarations

### Ethics approval and consent to participate

This retrospective study used exclusively irreversibly anonymised data generated during routine clinical care at Moulay Ismail Hospital, Meknes, Morocco. It was non-interventional and involved no modification of patient management. The protocol was reviewed by the Research Ethics Committee of the Faculty of Medicine, Euromed University of Fez, Morocco, which granted an exemption from full ethics review and waived the requirement for individual written informed consent, the anonymised nature of the dataset precluding re-identification of, and contact with, the patients concerned. The study was conducted in accordance with the principles of the Declaration of Helsinki.

### Consent for publication

Not applicable. The manuscript contains no individually identifiable patient data, images or clinical details permitting identification.

### Availability of data and materials

The anonymised datasets generated and analysed during the current study are not publicly available because of institutional restrictions on the sharing of patient-level clinical data, but are available from the corresponding author on reasonable request and subject to institutional approval.

## Author contributions

N.G. conceived and designed the study, collected and curated the data, performed the statistical analysis, interpreted the results, drafted the manuscript, and read and approved the final version.

## Data Availability

The anonymised dataset generated and analysed during the present study is not publicly available owing to institutional restrictions on the sharing of patient-level clinical data. It is available from the corresponding author on reasonable request and subject to institutional approval.

## Acknowledgements

The author thanks the clinical and nursing staff of the Department of Rheumatology, Moulay Ismail Hospital, Meknes, for their assistance with patient care and record retrieval.

## Clinical trial registration

Not applicable. This is a non-interventional retrospective observational study and was not registered in a clinical trial registry.

## Abbreviations

ACPA: anti-citrullinated protein antibody
ALT: alanine aminotransferase
AST: aspartate aminotransferase
CRP: C-reactive protein
csDMARD: conventional synthetic disease- modifying antirheumatic drug
DAS28: 28-joint Disease Activity Score
DMARD: disease- modifying antirheumatic drug
ESR: erythrocyte sedimentation rate
EULAR: European Alliance of Associations for Rheumatology
HDL: high-density lipoprotein
IL-6: interleukin- 6
LDL: low-density lipoprotein
MTX: methotrexate
RA: rheumatoid arthritis
RF: rheumatoid factor
SJC: swollen joint count
TCZ: tocilizumab
TG: triglycerides
TJC: tender joint count
TNF: tumour necrosis factor
ULN: upper limit of normal
VAS: visual analogue scale

## References

1. Guo Q, Wang Y, Xu D, Nossent J, Pavlos NJ, Xu J. Rheumatoid arthritis: pathological mechanisms and modern pharmacologic therapies. Bone Res. 2018;6:15. doi:10.1038/s41413-018-0016-9.

2. Smolen JS, Edwards CJ, Konzett V, et al. EULAR recommendations for the management of rheumatoid arthritis with synthetic and biological disease-modifying antirheumatic drugs: 2025 update. Ann Rheum Dis. 2026;85(6):991–1009. doi:10.1016/j.ard.2026.01.023.

3. Fraenkel L, Bathon JM, England BR, et al. 2021 American College of Rheumatology guideline for the treatment of rheumatoid arthritis. Arthritis Rheumatol. 2021;73(7):1108–1123. doi:10.1002/art.41752.

4. Rose-John S. Interleukin-6 signalling in health and disease. F1000Res. 2020;9:F1000 Faculty Rev- 1013. doi:10.12688/f1000research.26058.1.

5. Garbers C, Heink S, Korn T, Rose-John S. Interleukin-6: designing specific therapeutics for a complex cytokine. Nat Rev Drug Discov. 2018;17(6):395–412. doi:10.1038/nrd.2018.45.

6. Smolen JS, Beaulieu A, Rubbert-Roth A, et al. Effect of interleukin-6 receptor inhibition with tocilizumab in patients with rheumatoid arthritis (OPTION study): a double-blind, placebo-controlled, randomised trial. Lancet. 2008;371(9617):987–997. doi:10.1016/S0140-6736(08)60453-5.

7. Emery P, Keystone E, Tony HP, et al. IL-6 receptor inhibition with tocilizumab improves treatment outcomes in patients with rheumatoid arthritis refractory to anti-tumour necrosis factor biologicals: results from a 24-week multicentre randomised placebo-controlled trial. Ann Rheum Dis. 2008;67(11):1516–1523. doi:10.1136/ard.2008.092932.

8. Gabay C, Emery P, van Vollenhoven R, et al. Tocilizumab monotherapy versus adalimumab monotherapy for treatment of rheumatoid arthritis (ADACTA): a randomised, double-blind, controlled phase 4 trial. Lancet. 2013;381(9877):1541–1550. doi:10.1016/S0140-6736(13)60250-0.

9. Saki A, Rajaei E, Rahim F. Safety and efficacy of tocilizumab for rheumatoid arthritis: a systematic review and meta-analysis of clinical trial studies. Reumatologia. 2021;59(3):169–179. doi:10.5114/reum.2021.107026.

10. Behrens F, Burmester GR, Hofmann MW, et al. Sustained effectiveness and safety of subcutaneous tocilizumab over two years in the ARATA observational study. Clin Exp Rheumatol. 2023;41(7):1463–1472. doi:10.55563/clinexprheumatol/hlmsao.

11. Karadag O, Farisogullari B, Yagiz B, et al. Tocilizumab as a first line biologic agent in rheumatoid arthritis patients with inadequate response to disease-modifying anti-rheumatic drugs: real life experience from the TReasure Registry. Clin Exp Rheumatol. 2024;42(1):130–137.

12. Marsal Barril S, Martin-Martinez MA, Blanco-Garcia FJ, et al. Effectiveness and safety of tocilizumab in monotherapy in biologic-naive and non-naive patients with rheumatoid arthritis in a real-world setting. Reumatol Clin (Engl Ed). 2022;18(10):567–573. doi:10.1016/j.reumae.2021.12.004.

13. Jeon HL, Kim SC, Park SH, Shin JY. The risk of serious infection in rheumatoid arthritis patients receiving tocilizumab compared with tumor necrosis factor inhibitors in Korea. Semin Arthritis Rheum. 2021;51(5):989–995. doi:10.1016/j.semarthrit.2021.07.004.

14. Studenic P, Aletaha D, de Wit M, et al. American College of Rheumatology/EULAR remission criteria for rheumatoid arthritis: 2022 revision. Ann Rheum Dis. 2023;82(1):74–80. doi:10.1136/ard-2022-223413.

15. Pers YM, Fortunet C, Constant E, et al. Predictors of response and remission in a large cohort of rheumatoid arthritis patients treated with tocilizumab in clinical practice. Rheumatology (Oxford). 2014;53(1):76–84. doi:10.1093/rheumatology/ket301.

16. Forsblad-d’Elia H, Bengtsson K, Kristensen LE, Jacobsson LTH. Drug adherence, response and predictors thereof for tocilizumab in patients with rheumatoid arthritis: results from the Swedish biologics register. Rheumatology (Oxford). 2015;54(7):1186–1193. doi:10.1093/rheumatology/keu455.

